# Risk based tiered residential methamphetamine remediation limits

**DOI:** 10.1101/2025.03.23.25324483

**Authors:** J.E. Dennison

**Affiliations:** Century Environmental Hygiene, LLC, Fort Collins, CO 80524

**Keywords:** meth, risk assessment, exposure, dose, limit, standards

## Abstract

U.S. states that regulate methamphetamine (meth) contamination in residences require remediation to reduce meth surface concentrations to below a concentration limit. Existing limits vary widely, by 30-fold. States that have regulatory limits usually have a single limit that does not account for the extent of human contact with contaminated surfaces in different parts of residences (e.g., bedroom versus attic). Actual exposure in low contact areas does not contribute to total occupant dose as much as that in high contact areas, yet contributes similarly to remediation costs under the single limit paradigm. Heating, ventilation, and air-conditioning (HVAC) systems are also usually subject to the same limits despite low contact and without a basis in risk analysis. Methods for averaging test results into a compliance metric also affect remediation cost. This study analyzes the changes in total occupant meth dose and the concomitant reduction in remediation cost for various alternative concentration limits relative to these four issues. Residences were subdivided into normal occupancy areas (NOAs), low occupancy areas (LOAs) such as attics, moderate occupancy areas (MOAs) such as exterior hallways, and HVAC systems. Dose calculations were based on existing risk assessment methodologies and cost reductions were based on contractor survey data and random residence sampling data. Raising only the conservative remediation limits adopted by some states to the California single limit would reduce the average cost to remediate a contaminated house from $21,000 to $10,000. Further raising limits to concentrations that remain below risk assessment boundaries is estimated to save an additional $5,000 on LOA and $2,000 on HVAC per remedial project. If all recommended alternate concentration limits are implemented, an estimated 50% additional cost reduction is possible with tiered limits based on contact time. These alternate limits still allow the occupant dose to fall below the recommended dose levels, which implies that these LOAs, MOAs, and HVAC systems are being over remediated when they are required to be remediated to the same limit as NOAs.

## INTRODUCTION

Meth contamination in residences is a significant on-going problem in the U.S. and in other countries where meth use occurs. The average cost to remediate a property, including associated repair costs, was estimated at $44K and over 5 million U.S. residences, 4% of U.S. housing stock, were estimated to be contaminated above a remediation standard of 0.5 μg/100 cm^2^, occupied by 13 million people (Dennison 2025). Exposure to non-users could be secondhand during meth manufacturing or smoking activity, or more often, thirdhand, after activity ceases. Thirdhand exposure occurs because meth contamination persists for extended periods as surface residues and particulates in indoor spaces (Bitter 2017). Approximately 99% of contamination is from meth use via smoking, not manufacturing. With most states having use rates within twofold of the national average use rate, contaminated residences are ubiquitous (Dennison 2025). Yet, only 19 states currently regulate meth contamination in residential properties, and of these states, 14 require remediation only when there is evidence of manufacturing, not when only use occurred (MLCC 2024) despite the prevalence of use instead of cooking. One of the most probable reasons for this regulatory gap is the high cost of remediation.

In general, existing meth regulations specify that residences with meth activity that is subject to the rule must be tested, and areas that exceed a surface concentration limit (μg meth/100 cm^2^) must be remediated. The limit is a surface concentration, as meth risk assessments have concluded that exposure is mainly through dermal and ingestion pathways and not inhalation (CalEPAa 2009, Hammon and Griffin 2007). Seventeen states have a single limit that applies to all parts of the property (MLCC 2024). The absorbed meth dose is associated with both the 1) concentration of meth residues and 2) the length of exposure and areal extent of contact with contaminated surfaces. Thus, parts of buildings where occupancy is limited, low occupancy areas (LOAs) and moderate occupancy areas (MOAs), contribute less to the total dose. Two states (Colorado and Wyoming) have promulgated a novel approach by developing two-tier limits with higher limits for LOAs. Wyoming set a limit of 3.75 μg/100 cm^2^ for attics, crawlspaces, and heating, ventilation, and air-conditioning (HVAC) systems (Wyoming 2010) versus a limit of 0.75 μg/100 cm^2^ for the rest of the residence. Colorado established a limit of 4 μg/100 cm^2^ for attics and crawlspaces and similar areas versus 0.5 μg/100 cm^2^ for other areas.

In the 19 states that set limits, the range is 30-fold, from 0.05 to 1.5 μg/100 cm^2^ (MLCC 2024). Compared with residential radon, asbestos and lead standards, which vary little from state to state, this is a wide range. Part of the reason for is that some states set limits at the minimum detectable level prior to the publication of meth risk assessments. After Colorado and California published their risk assessments with recommended limits of 0.5 μg/100 cm^2^ and 1.5 μg/100 cm^2^ (CDPHE 2005; CalEPA 2009b, 2009a), respectively, several states promulgated limits in that range (Wyoming 2010) or raised their lower limit to the same range (Washington 2024), while many other states did not modify their limits.

All states except Wyoming currently regulate contamination in HVAC systems similarly to dermally accessible surfaces such as countertops or walls. The dermal and ingestion dose calculations used for accessible surfaces do not apply to contaminated HVAC systems and a different paradigm is needed. Another issue relates to the regulatory requirement that every sample within a room or space, and every sample within different rooms or spaces must meet the regulatory limit. Exposure to occupants depends on concentrations on contact surfaces, and the total dose recommended in the meth risk assessment would not be exceeded if the average concentration contacted is less than the meth limit. As room concentrations vary, the average concentration in a building where every room meets the limit will be less than the limit, resulting in a total dose that is less than required. Thus, several inconsistencies or questions arise regarding how meth limits have been set in various jurisdictions and the impact of applying single limits to all space types, and of not allowing whole-building averaging. The purpose of this study is to provide a methodology for calculating the incremental dose when varying meth limits are applied to different space types. Using this methodology, the study provides calculations of the increased dose for several tiers of alternate meth limits (AMLs).

There would be no purpose for considering higher remediation limits were there not concomitant cost savings, so this study also examines the impact of AMLs on the number of buildings that require remediation and the associated costs. Remediation costs are tied to remediation limits in several ways. First, fewer properties would exceed a higher limit and require remediation, and a smaller portion of properties would exceed a higher limit (an entire house may exceed a lower limit and only half the house may exceed a higher limit). Second, it is incrementally more difficult to reduce contaminant levels to lower limits, so the cleanup cost per square foot is less when higher limits apply. Thus, this study will provide the cost reduction estimates alongside the dose calculations.

## METHODS

### Incremental Dose Calculations

Two health risk assessments have been published for thirdhand exposure to methamphetamine (Hammon and Griffin 2007, CalEPA 2009a,b). Both risk assessments determined that exposure was via dermal and ingestion routes. The dermal dose was calculated using equations of the form:

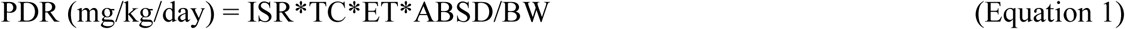

where PDR = potential dose rate (mg/kg/day), ISR = indoor surface transferable residue (mg/cm^2^), TC = transfer coefficient (cm^2^/h), ET = exposure time (h/day), ABSD = dermal absorption fraction (unitless), and BW = body weight. Ingestion dose was calculated using a substantially similar equation, and total dose was the sum of dermal and ingestion doses. The dose rate is proportional to all parameters including the ISR and ET. The dose for any scenario is calculated by inserting either the existing limit (the base meth limit (BML) or AML for ISR. The incremental dose would be the percent increase in dose from:

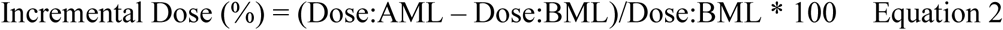

Where the other parameters (TC, ABSD, and BW) would appear in Eq.2, they cancel out and the incremental dose can be simply calculated according to Equation 3, which allows different limits for different space types:

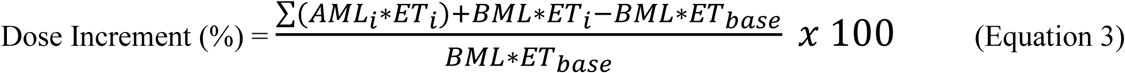

where AML*_i_*= alternate meth limit (μg/100 cm^2^), BML = base meth limit (μg/100 cm^2^), and ET = exposure time (hrs), each for space type *i*.

LOAs were defined to include attics, crawlspaces, and analogous spaces that are not intended or used for purposes other than storage or utilities. MOAs were defined as garages, outbuildings, porches, interior hallways, exterior balconies, and exterior stairwells. The average amounts of time occupants spend in LOAs and MOAs was assumed to be 10 hrs./year for LOAs and 1 hr./day for LOAs (Table 1). The BML used for dose calculations was the Colorado standard (0.5 μg/100 cm^2^).

**Table 1:**
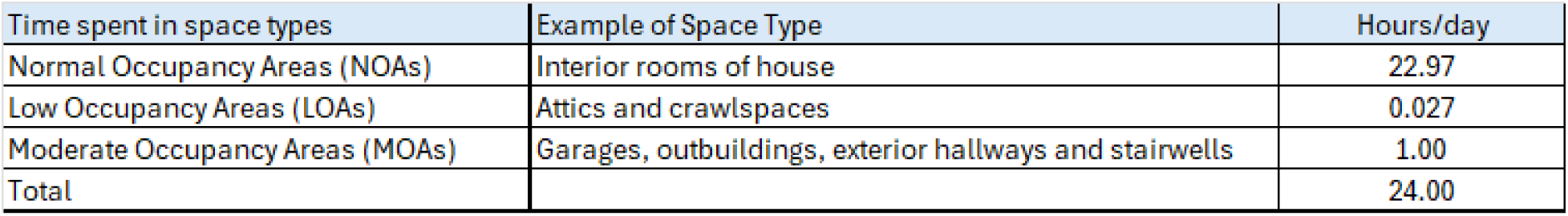
Estimate of time spent in NOAs, LOAs, and MOAs.

### Building Contamination and Remediation Cost Model

In order to estimate remediation costs under different meth limits, contaminant data from representative buildings was needed. Comprehensive tests normally report contaminant concentrations for different rooms, areas, and systems in a residence, hereafter referred to as a Housing Unit (HU). These data allowed determination of specific rooms and areas that exceed the BML or AML being considered and the associated costs for any limit scenario. The data for contamination analysis consisted of 100 Preliminary Assessment (PA) reports randomly selected from the Colorado Department of Public Health and Environment (CDPHE) online repository of PAs (CDPHE 2024). The 100 HUs consisted of 1-story apartments (21), 2–4-unit multiplexes (13; one unit each), single-family homes (52), and townhomes (14). In a PA, one four-part composite wipe sample is collected in each room and four discrete samples in HVAC, and each room or area is designated as contaminated or not based on the result for each sample (and highest sample in HVAC). The PAs were conducted by industrial hygienists at 10 certified meth consulting firms in Colorado. All samples were analyzed by accredited laboratories using either NIOSH 9109 (Methamphetamine and Illicit Drugs, Precursors, and Adulterants on Wipes by Solid Phase Extraction) or NIOSH 9111 (Methamphetamine on Wipes by Liquid Chromatography/Mass Spectrometry). Three reports were rejected and replaced with another due to report errors, and all the reports from one consultant were omitted as the consultant collected HVAC samples primarily on the exterior of ductwork and furnace housings, not in the airflow pathway. The results for each room, area or space were then tabularized. For the purposes of this analysis, “HVAC systems” refers to central heating, ventilating, and air-conditioning systems that have a heater (usually a furnace), a fan to move air through supply ductwork, return ducts (which may be very short in smaller apartments), and possibly cooling equipment. Non-ventilating hydronic systems, wall unit heaters, air conditioners, or other heating and cooling systems that lack ductwork were not included in this analysis. Auxiliary local exhaust systems such as bathroom exhaust fans or whole house fans are addressed separately. In the dataset used here, 84% of the HUs had central HVAC. Contaminated HVAC means the meth concentration inside the ducts and equipment, not on the exterior surface. Single-point (discrete) samples were usually provided for different parts of HVAC systems, and up to two samples were collected in supply and return ducts. In some instances, multi-point composite samples from HVACs were reported and coded as a single sample under “furnace.” Dose calculations for dermal/oral exposure to interior HVAC contamination were based on assumed contact time of one hour per year, a worst-case estimate of time required for four filter changes.

Non-house “rooms” included attics, crawlspaces (which may be below the house or of the cubbyhole type), garages, sheds, etc. Redundant rooms were listed separately (e.g. Bedroom 1, 2…) and unique room types were coded as a miscellaneous room. Data for local exhaust systems and separate air conditioners were also coded if available. Samples from contents (personal property) and appliances were not included as they are only sampled sporadically and are usually disposed of, the cost thereof built into the base remediation cost. If data were reported as “Below Reporting Level,” the Reporting Levels were used. Sample photographs and drawings were reviewed to confirm locations when sample location descriptions were unclear. Limited report reviews were conducted, primarily to verify that samples noted as HVAC and bathroom fans were collected inside the system. As painting over meth can have a significant effect on results, if painting prior to the PA was noted, the data were flagged. For this analysis, the area failed if the result was greater than the limit. Concentration data from hallways in 12 apartment buildings with a contaminated unit were also compiled. Data was also extracted from 15 post-remediation reports randomly obtained from the CDPHE online archive (CDPHE 2024).

A sample record is provided in Table 2. The Meth Concentration in Rooms/Areas detail the PA results for the HU. The Concentration Summary lists the highest or average result for each space type. The Logic Functions section details whether each space type passed or failed by comparing the highest result for the space type with the limit. Based on the cost model, the following space types were needed: rooms (and # of rooms that failed), HVAC, MOAs, attic, and crawlspace. Remediation cost data were collected from contractors who are certified or specialized in meth remediation in the U.S. Publicly available lists of meth contractors were obtained from the internet, and all listed contractors were sent questionnaires. Fifteen responses were received and averaged (Table 3). There were obvious differences in costs in several states, which may partly depend on state regulations, such as numeric remediation limits and the process for remedial verification, as well as local cost conditions. The table provides the average remediation cost for a 10-room house as well as additional costs for LOAs and MOAs and costs for additional or fewer than 10 rooms. There were many variables taken into consideration for estimating remediation costs, so these costs were strictly an average. These data were then used to estimate the remediation cost for each contaminated HU based on all rooms and areas that failed. For the analysis of remediation costs with AMLs, the spreadsheet code calculated the overall cost for remediation of each HU based on the limits applied.

**Table 2:**
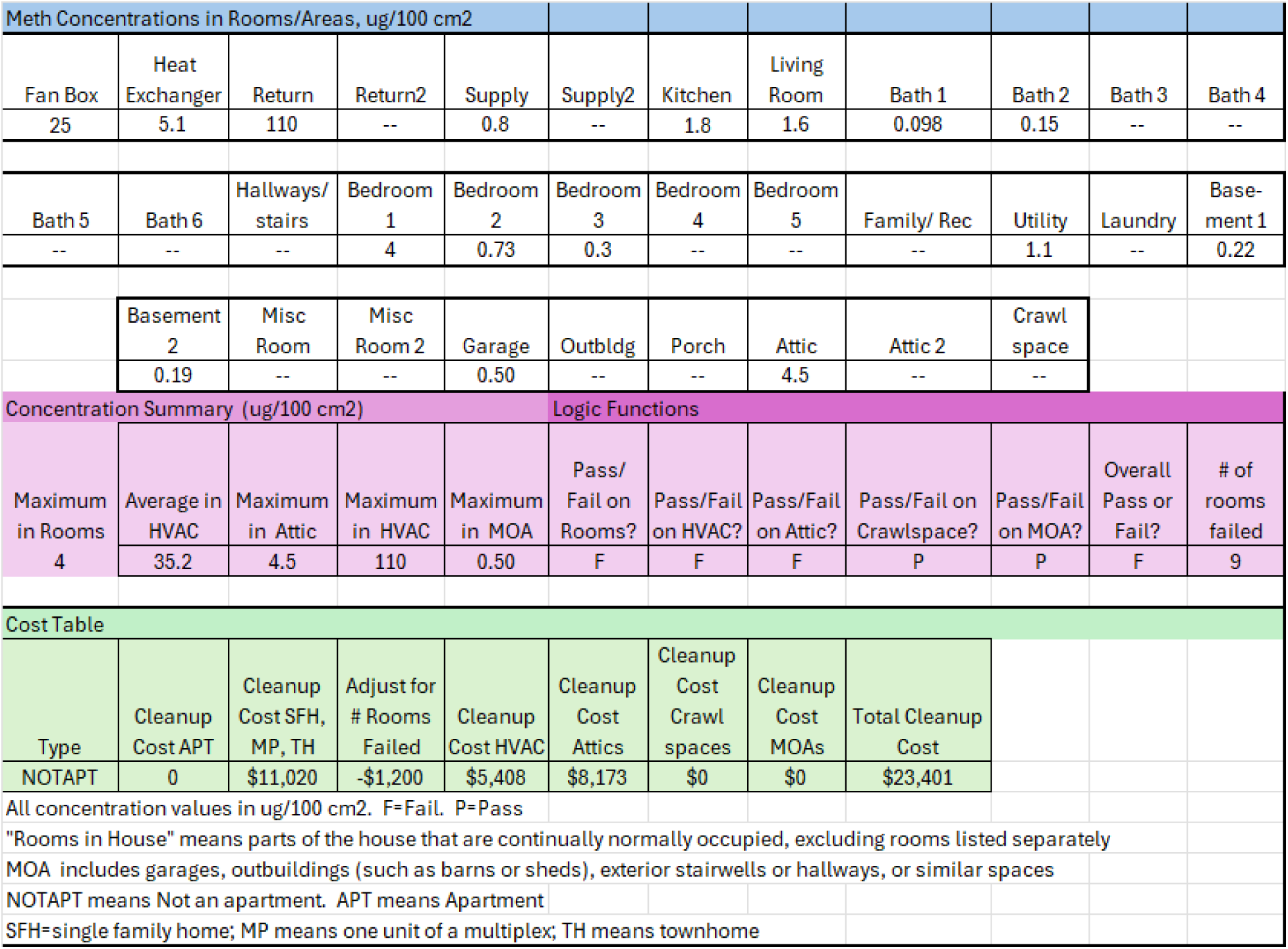
Concentration and cost model.

**Table 3:** Average remediation costs.

| Remediation Costs (U.S. Average) | Cost (US\$) |
| --- | --- |
| Apartment, 2 bedroom 1 bath | \$6,088 |
| SFH ranch style house, 3 bedroom, 3 bath, attached garage | \$11,020 |
| Additional cost if HVAC is contaminated | \$5,408 |
| Additional cost if an attic is contaminated | \$8,173 |
| Additional cost if a crawlspace is contaminated | \$7,436 |
| Additional cost if garage, outbuildings, porches etc contaminated | \$1,657 |
| Adjust per additional/fewer rooms | \$1,200 |

No repair costs are included in the remediation estimates except as noted next. The cleaning costs listed in Table 3 include additional cleaning for areas that do not pass a post-remediation sample. Meth can be difficult to remediate, and many properties require several cleanings until each area passes. HVAC systems are frequently one of the most difficult areas to clean. In many instances, the furnace and even the supply or return ducts may be removed rather than cleaned or re-cleaned. From the author’s experience, the furnace is removed ∼40% of the time (especially if it approaches a 20-year service life) and ducts are removed on 15% of projects. As replacements costs are substantial, the costs for new furnaces ($4,000) and ducts ($7,500) were factored into the HVAC mitigation cost pro-rata based on average system costs (HomeGuide 2024). Likewise, if an attic is remediated, insulation is usually removed, and if a crawlspace is remediated, a vapor barrier may be removed and/or installed. Thus, these costs were added for attic insulation at $2,500 (Bob Villa 2024) and vapor barriers at $2,600 each (RFR 2024). Ancillary costs such as replacing drywall, carpet, appliances, fixtures, cabinets, post-remedial testing; loss of use; and legal expenses are difficult to estimate in a manner that would apply to all projects, so were excluded. Thus, the costs provided here are a minimum.

While there can be more than one attic space and occasionally more than one crawlspace, the average cleanup cost is applied if any attic or crawlspace exceeded the remediation limit. MOAs were similarly grouped, and the estimate included the line-item cost if any MOA was deemed contaminated. The cost to remediate all areas that failed were totaled to obtain a cleanup cost for that HU, and then the average of cleanup costs for all properties HU that failed the AML was calculated. The number of HUs contaminated per year in the U.S. was based on 2022 data (Dennison 2025). The spreadsheet (available in supplementary data) can be revised with state-specific contamination and cost data and re-calculated for any combination of BMLs and AMLs.

## RESULTS

The average concentrations of meth in all room and space types for the 100 assessments used in this study are shown in Table 4. Components of the HVAC system and bathroom vents typically had the highest concentrations, and garages, outbuildings, and porches were among the lowest. For the BML of 0.5 μg/100 cm^2^ applied to all areas, 86% of HUs were deemed contaminated. Failure rates in NOAs, LOAs, MOAs, and HVAC were 71%, 43%, 21% and 68%, respectively. Post-decontamination results found average concentrations in remediated houses to be 0.1 μg/100 cm^2^ in the NOA of HUs and 0.15 μg/100 cm^2^ in HVAC systems.

**Table 4:** Meth concentrations in 100 HUs.

| Space Type | Average,<br>ug/100 cm2 | Standard<br>Deviation | Normalized |
| --- | --- | --- | --- |
| Outbuildings | 0.2 | 0.4 | 1 |
| Porch | 0.6 | 1.9 | 3 |
| Garage | 0.9 | 1.8 | 4 |
| Crawlspace | 1.3 | 2.0 | 6 |
| Family/ Rec Room | 1.5 | 2.5 | 7 |
| Bathrooms | 2.1 | 6.2 | 10 |
| Bedrooms | 2.1 | 8.9 | 10 |
| Hallways/stairs | 2.1 | 4.1 | 10 |
| Other Vents | 2.2 | N/A | 11 |
| Attic | 3.1 | 5.5 | 15 |
| Miscellaneous Room | 7.0 | 29 | 34 |
| Kitchen Vent | 7.0 | 14 | 34 |
| Living Room | 10.0 | 57 | 48 |
| Kitchen | 11.3 | 52 | 54 |
| Supply Ducts | 11.3 | 30 | 54 |
| Laundry Vent | 11.5 | 12 | 55 |
| Laundry | 12.1 | 54 | 58 |
| Utility | 16.7 | 66 | 80 |
| Basement | 19.0 | 85 | 91 |
| Furnace Fan Chamber | 23.2 | 49 | 112 |
| Furnace Heat Exchanger | 26.6 | 93 | 128 |
| Return Ducts | 62.9 | 226 | 303 |
| Air Conditioner | 71.5 | 75 | 344 |
| Bathroom Vents | 298.0 | 1386 | 1434 |

### Conservative Remediation Limits

Figure 1a shows the percent of HUs that fail and the associated remediation cost for BMLs between 0.05 and 10 μg/100 cm^2^ using no AMLs (only single tier limits). At the lowest extant limit of 0.05 μg/100 cm^2^, 98% of the HUs required remediation, and this declined to 86% at 0.5 μg/100 cm^2^ and 70% at 1.5 μg/100 cm^2^. Per project remediation costs were based on the failure rates for BMLs of 0.05 and 0.1 μg/100 cm^2^ (98% and 97% respectively) but were based on the 86% failure rate for all other limits, as using each BML failure rate would not account for cost savings from HUs that would pass at higher limits. The cost per remediation project decreased from $21K at 0.05 μg/100 cm^2^, to $15K at 0.5 μg/100 cm^2^, and $10K at 1.5 μg/100 cm^2^ and decreased further at higher limits. Figure 1b shows the specific cost reduction per remediation project for different permutations of original and higher limits. For example, if a regulation specified a current limit of 0.1 μg/100 cm^2^ and raised the limit to 1.5 μg/100 cm^2^, a cost reduction of $9,600 per project would be realized. Dose decrements would be proportional to the ratio of any BML as compared to either Colorado or California limits. For example, a BML of 0.05 μg/100 cm^2^ would result in 10% of the dose received under the Colorado limit.

**Figure 1a:**
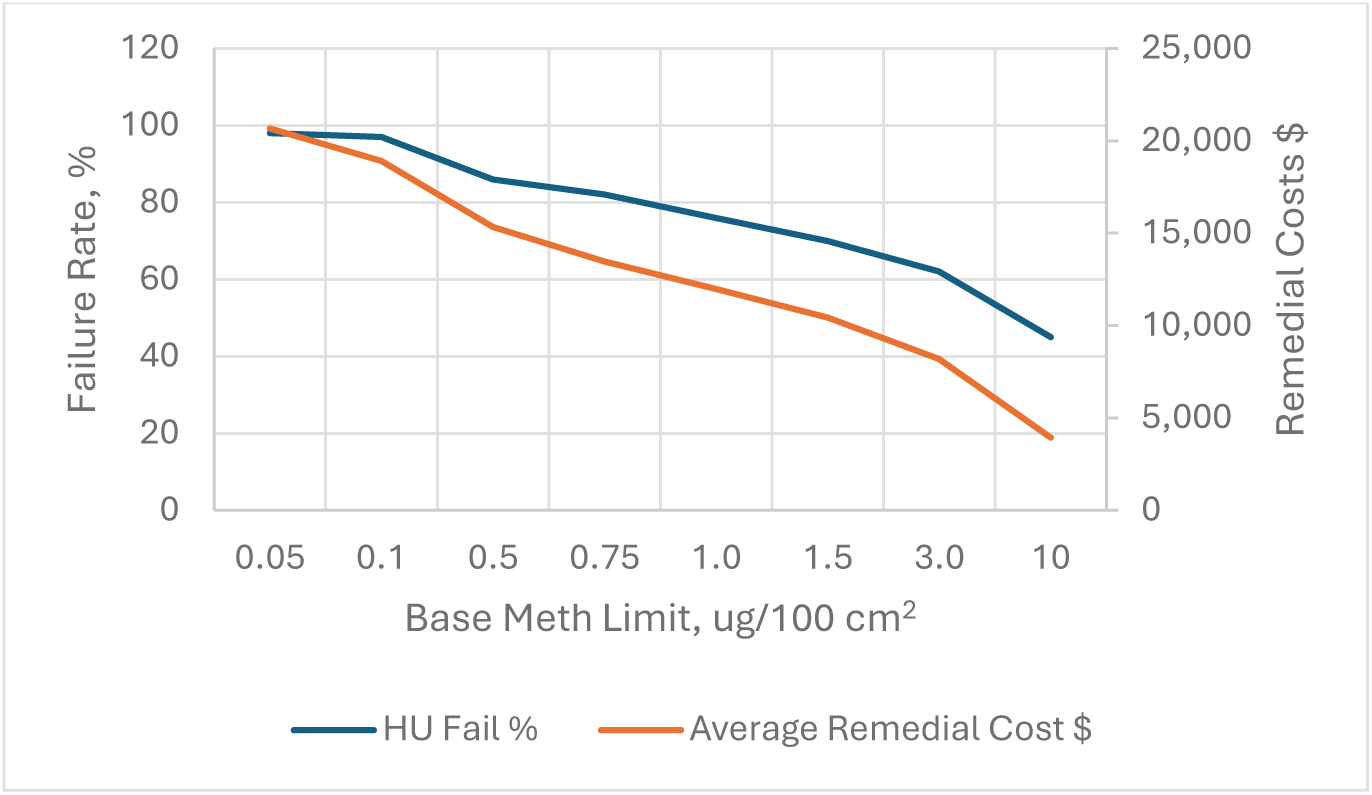
Failure frequency and remediation costs for different single remediation limits.

**Figure 1b:**
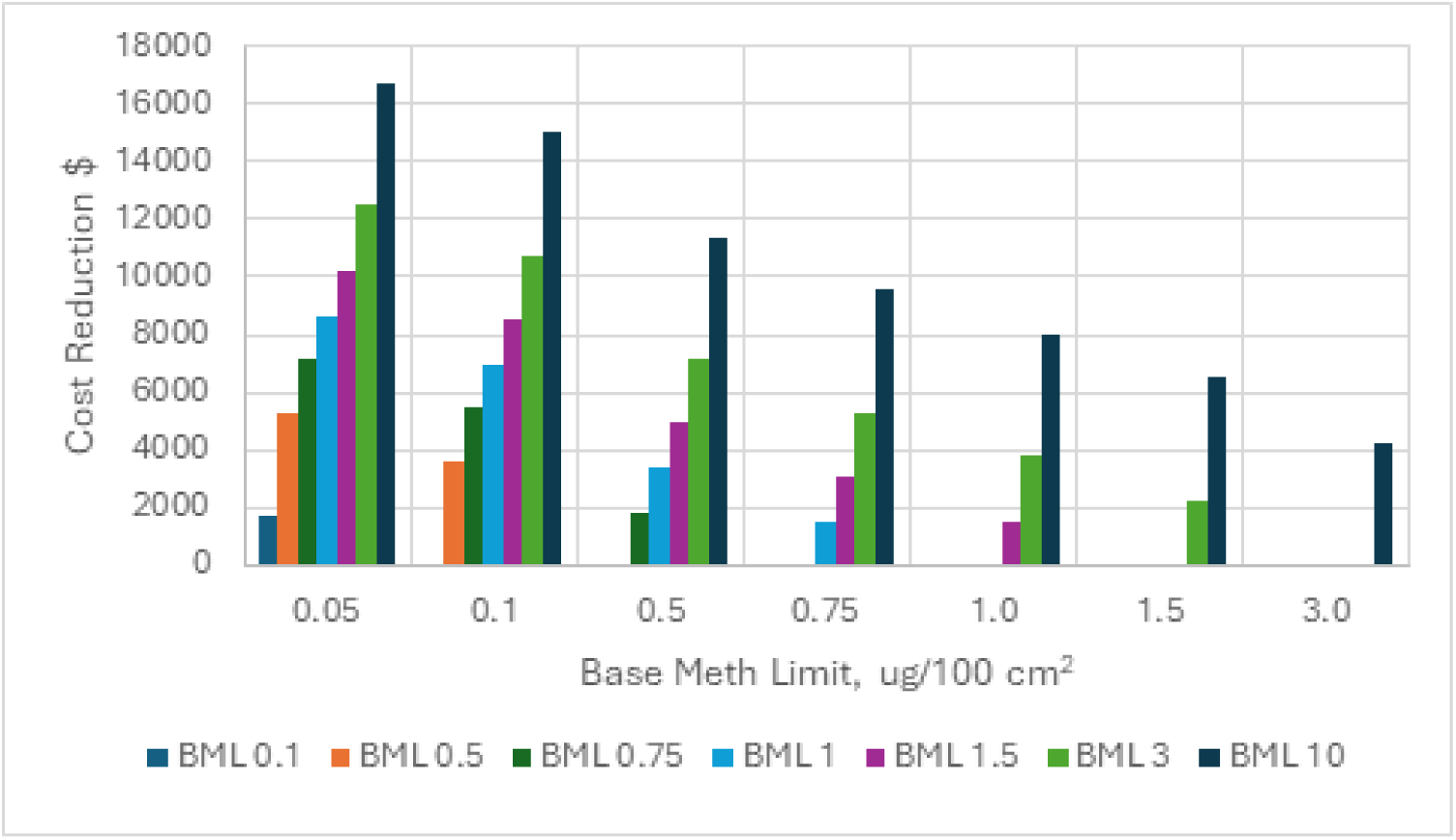
Cost reductions associated with increasing remediation limits.

### Low And Moderate Occupancy Areas

For the BML of 0.5 μg/100 cm^2^ and an AML for LOAs of 4 μg/100 cm^2^, the incremental dose is 0.8%. and is 2.2% at 10 μg/100 cm^2^ (Figure 2b). The same incremental doses would apply to any BML. For example, if the BML was 1.5 μg/100 cm^2^, an eight-fold increase of the LOA limit would also result in a 0.8% increase to total dose. Results of the LOA cost calculations are shown in Figures 2a and 2b. The calculations use a BML of 0.5 μg/100 cm^2^ and a failure percentage of 86%. Remedial costs decline ∼$4K and $5K per project when the LOA limit is set to 4 or 10 μg/100 cm^2^, respectively. Further reductions with yet higher LOA limits are modest as at 4 μg/100 cm^2^, only 4% of crawlspaces and 15% of attics fail. The overall HU failure rate is not affected very much because other parts of the HUs still fail.

**Figure 2a:**
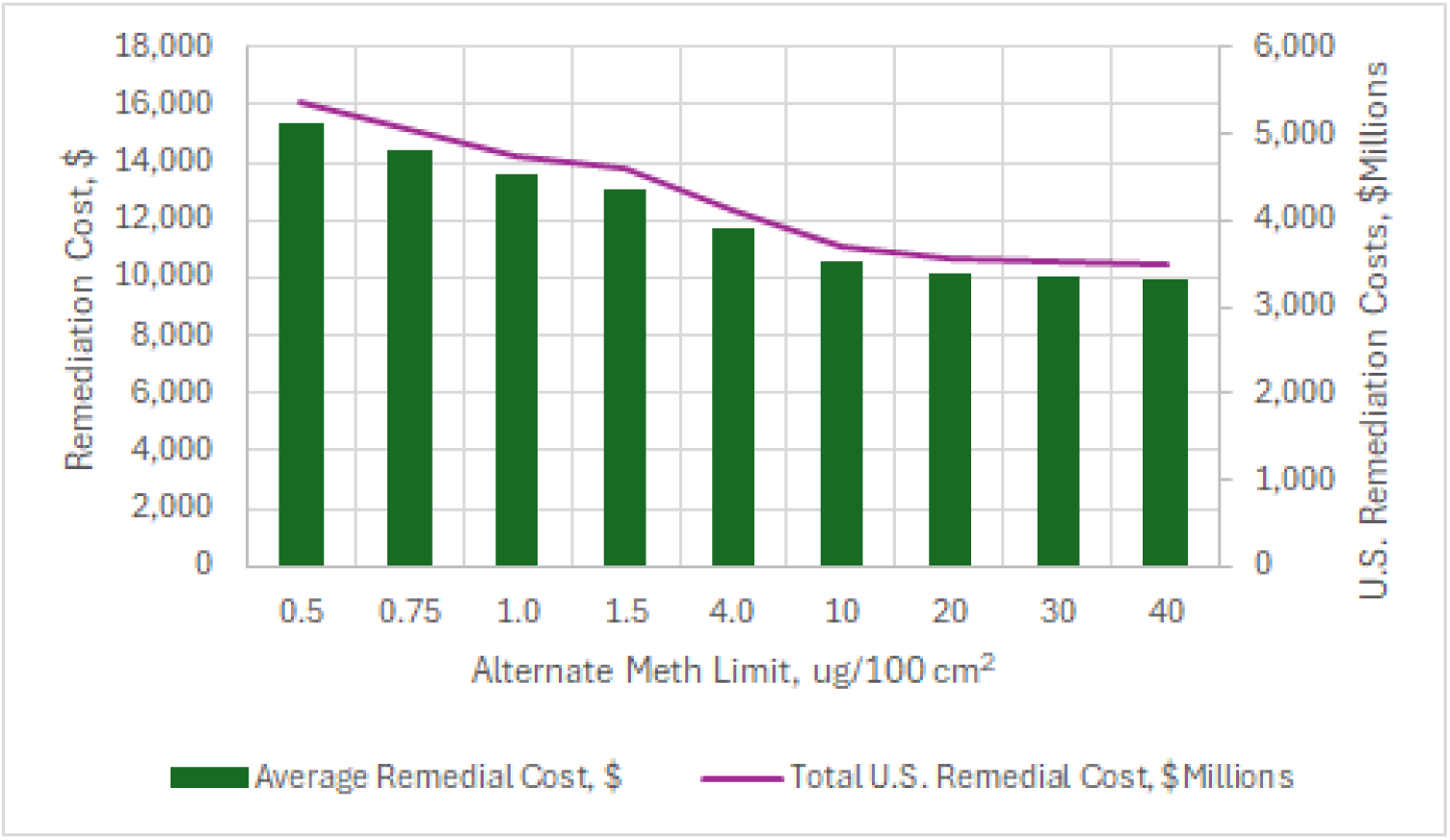
Average remedial costs per project and total U.S. remedial costs for LOA AMLs

**Figure 2b:**
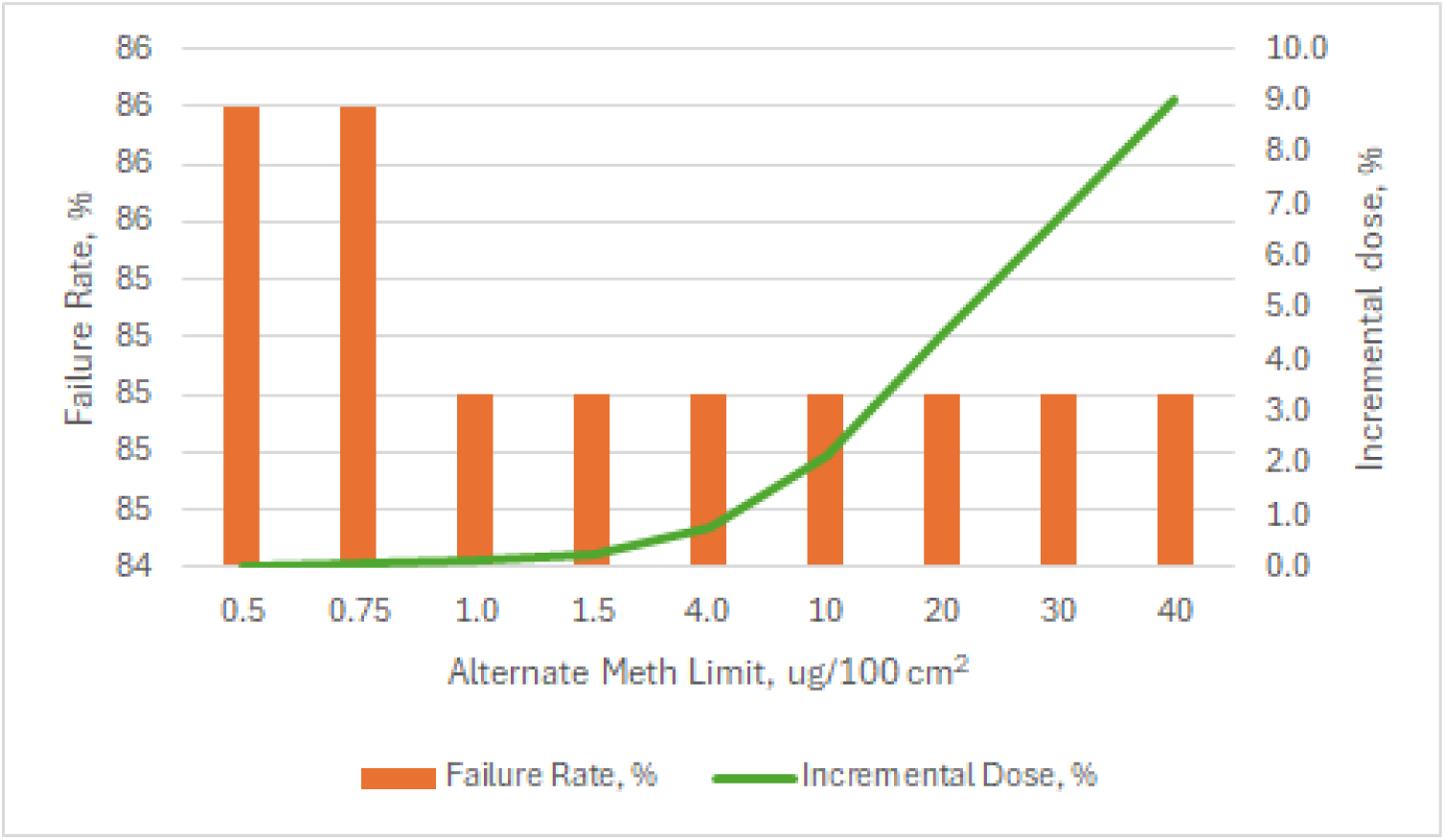
Percent of HUs that fail and incremental dose calculations for LOA AMLs

The same analysis was conducted for MOAs, which were assumed to be occupied for one hour per day. MOAs primarily consist of garages and outbuildings in single family homes, which are not a focus of this study, and hallways and stairwells in multifamily buildings, which are the main concern. The incremental dose is 4%, 8% and 29% at 1, 1.5 and 4 μg/100 cm^2^, respectively. Data in hallways adjacent to contaminated units indicated that hallways exceeded 0.5 μg/100 cm^2^ 33% of the time and exceeded 1 μg/100 cm^2^ 17% of the time. The ratio of meth concentrations in the source units’ adjacent room to hallways was 76:1. Modest decreases in remedial costs per project and in failure rates are found after varying MOA AMLs from 0.75 to 10 μg/100 cm^2^ (Figures 3a and 3b), where the failure rate for MOAs reached 0%.

**Figure 3a:**
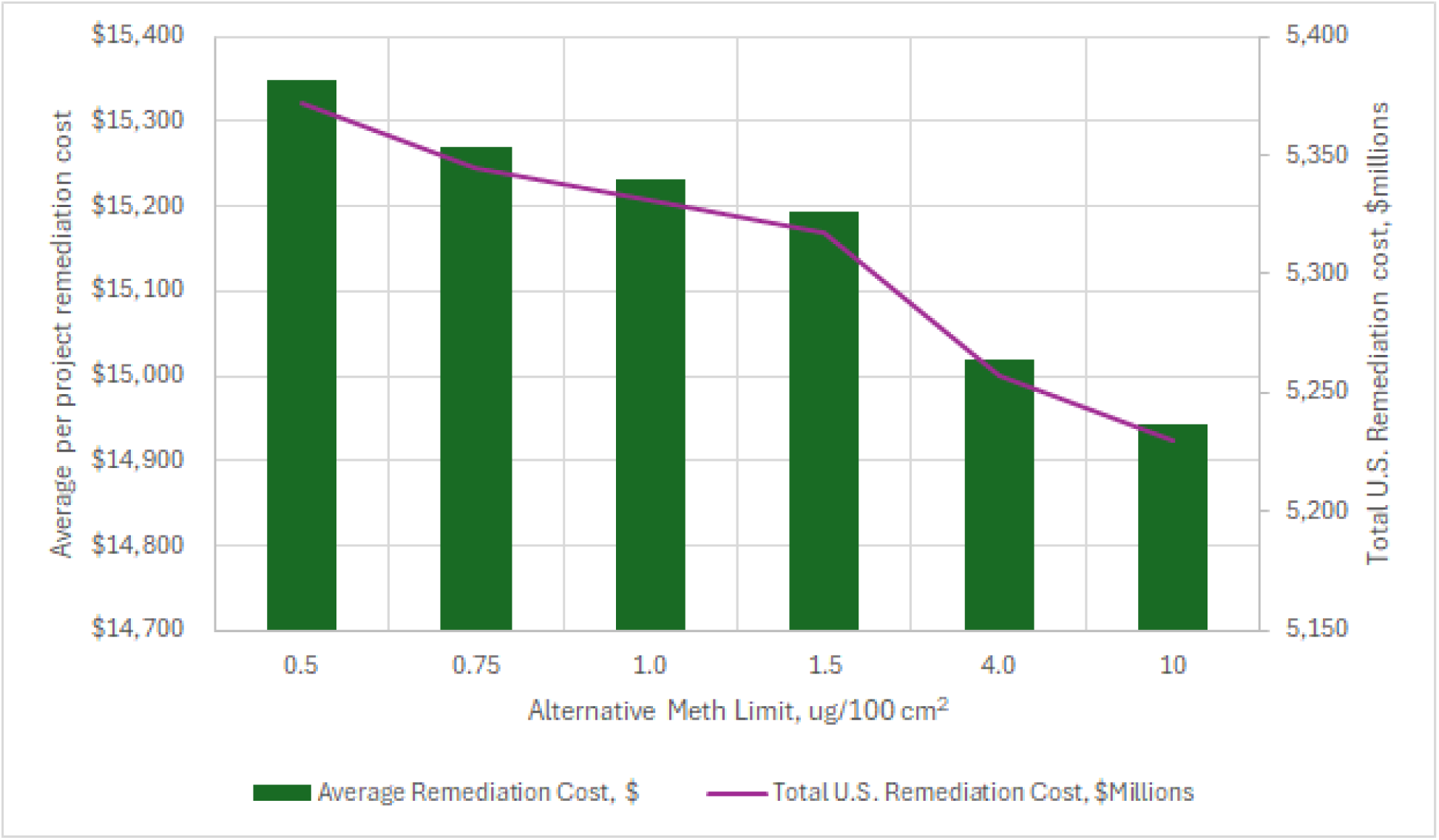
Average remediation costs per project and total U.S. remediation costs for AMLs for MOAs

**Figure 3b:**
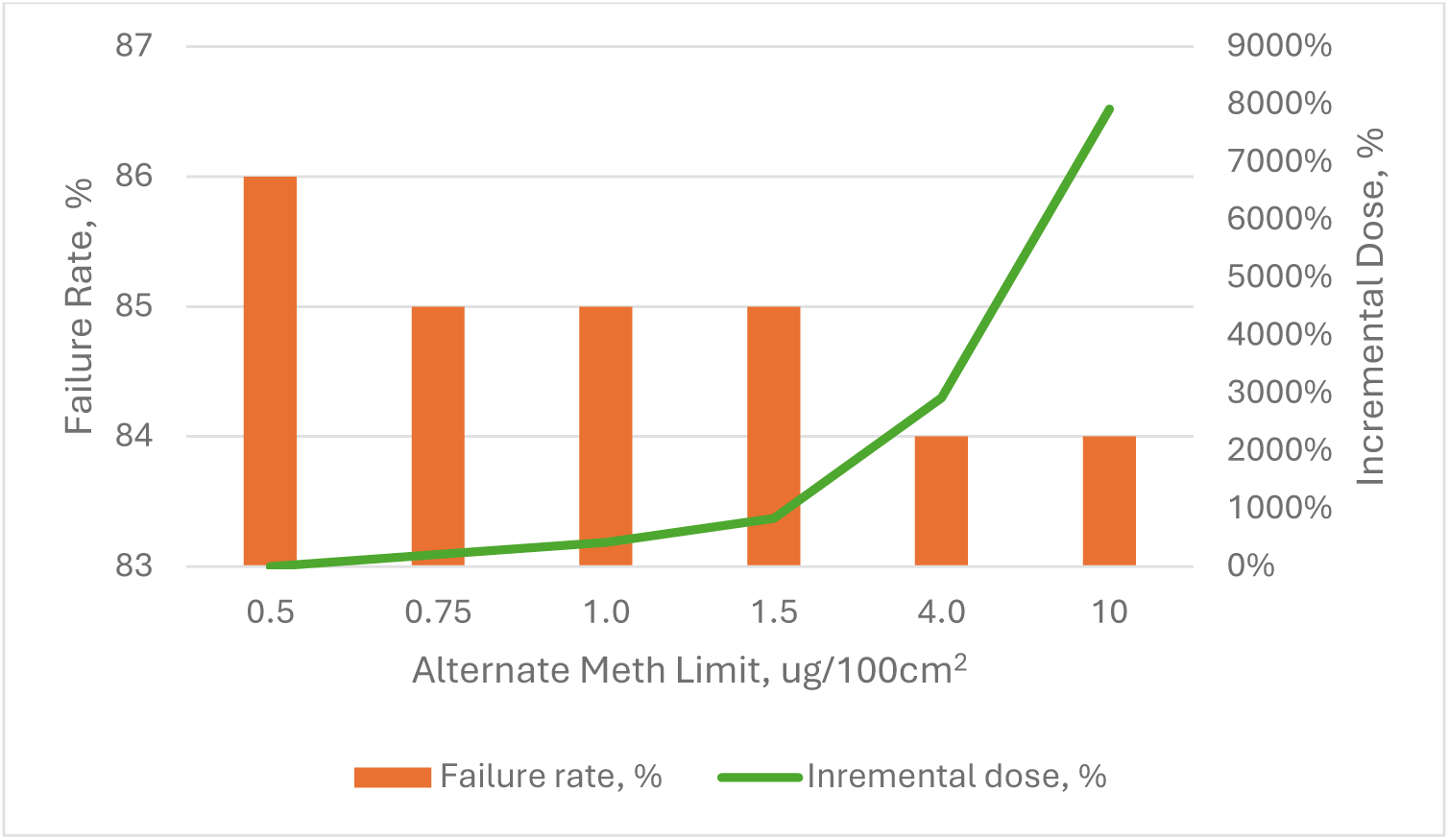
Percent of HUs that fail and incremental dose calculations for AMLs in MOAs

### HVAC Systems

The incremental dermal/oral dose for a limit of 10 μg/100 cm^2^ is 0.1%. The cost of remediating HVAC systems to the BML and AML and the resultant failure rates are shown in Figure 4. These data indicate that, for a BML of 0.5 μg/100, increasing the HVAC remediation limit to 4 or 10 μg/100 cm^2^ would reduce costs by ∼$900 and ∼$1,800, respectively, per remediation project if the HVAC maximum discrete sample concentrations are used. The average concentration in post-remediation samples was 0.15 μg/100 cm^2^.

**Figure 4:**
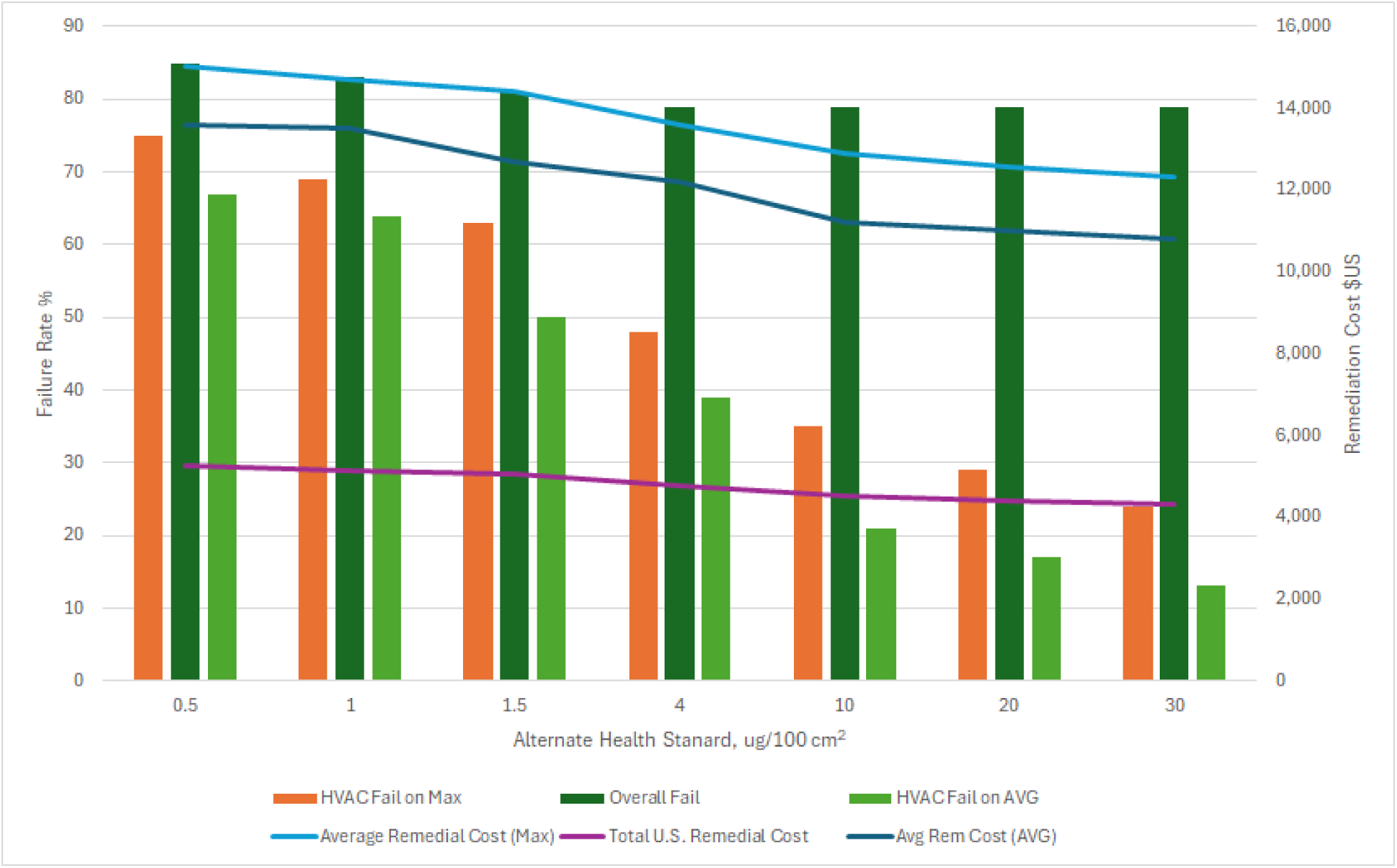
HVAC Remediation costs and failure percentages using the maximum discrete sample result (MAX) and averaged (AVG) discrete sample results

### Combined AMLS

Different AMLs can be simultaneously promulgated for NOAs, LOAs, MOAs, and HVACs. The combined assessment for these recommended AMLs appears in Table 5 with respect to BMLs of 0.5 and 1.5 μg/100 cm^2^. The LOA and HVAC AML levels were varied up to 20-fold of the BML and the MOA AML was varied up to 1.5 μg/100 cm^2^. Relative to the BML of 0.5 μg/100 cm^2^, a cost reduction of 44% was obtained if the MOA limit is 1.5 μg/100 cm^2^ and the HVAC and LOA limit was raised to 10 μg/100 cm^2^. The estimated average remediation cost for a remediation limit of 1.5 μg/100 cm^2^ was $10K and was reduced by 50% if the MOA limit is 1.5 μg/100 cm^2^ and the HVAC and LOA limit was raised to 30 μg/100 cm^2^.

**Table 5:** Failure rates and remediation costs for increased AMLs.

| AML (LOA/HVAC),<br>$\mu\text{g}/100\text{ cm}^2$ | Overall Fail % | Average Remedial Cost (\$) | Cost Reduction (\$) |
| --- | --- | --- | --- |
| Base Limit 0.5 $\mu\text{g}/100\text{ cm}^2$ | | | |
| 0.5 | 86 | 15,328 |  |
| 1 | 82 | 13,397 | 1,931 |
| 1.5 | 80 | 12,316 | 3,012 |
| 5 | 75 | 10,035 | 5,293 |
| 10 | 72 | 8,600 | 6,728 |
| Base limit 1.5 $\mu\text{g}/100\text{ cm}^2$ | | | |
| 1.5 | 70 | 10,423 |  |
| 7.5 | 59 | 7,236 | 3,188 |
| 15 | 58 | 5,950 | 4,473 |
| 30 | 57 | 5,226 | 5,197 |

## DISCUSSION

### Regulatory Gap Analysis

Twelve states regulate meth only after cooking and not after use, although in some localities if a property tests positive, remediation may be required through local codes or similar mechanisms. Currently, 31 states have no public health regulations concerning meth in residences. An estimated 4% of all U.S. housing is currently contaminated with meth at a concentration above 0.5 μg/100 cm^2^ and ∼99% of newly contaminated housing is contaminated by use rather than cooking (Dennison 2025). As meth health risk is not based on the type of activity that caused contamination to exist, but based on the presence of meth alone, these are regulatory gaps.

### LOAs

As previously discussed, the dose received by occupants is proportional to the amount of time an occupant spends in a room or area. Children or other occupants may spend time and have contact with contaminated surfaces in various rooms of the house, but a much smaller proportion of time is spent in LOAs and MOAs, such as attics. If the limit does not account for occupancy time, it is implicitly assuming that the amount of time spent in a space such as an attic is 1/*n*^th^ of the amount of time spent in NOAs, where *n* is the number of rooms in the house +1. So, if the HU is a six-room house with an attic space, the single limit approach assumes that the occupant could spend 1/7^th^ of their time in the attic (∼3.5 hrs/day) to receive an equal fraction of total dose from all spaces. Of the 19 states that have regulatory limits, all but two set one remediation limit for all parts of a property. Thus, all rooms that are tested, including HVAC systems, attics, or other spaces, must meet the single limit or be remediated to that limit. If a limit for LOAs were increased from 0.5 μg/100 cm^2^ to 4 μg/100 cm^2^, the incremental dose is 0.8% and is 2.2% for a 10 μg/100 cm^2^ limit. Eight and twenty-fold increases with respect to other state limits would result in the same nominal dose increments. Cost savings of ∼4K or more per project are worth consideration due to this nominal effect on total dose.

### MOAs

The estimated time for MOA occupancy was 1 hr/day, which may greatly exceed the average time spent coming and going in hallways or stairwells, but may be experienced by some occupants in a garage or other MOA. While this may potentially underestimate exposure time for a fraction of adults, it may reasonably estimate maximum exposure time for young children, for which the remediation limits were established. Colorado published an adult exposure limit of 4 μg/100 cm^2^ (Goyal 2014), so the exposure time for adults is not necessarily an issue.

Only modest cost savings appear possible if MOA limits were raised to 1, 4 or even 10 μg/100 cm^2^. However, this assessment ignores important additional costs that are difficult to quantify but are easily described. From an industrial hygiene standpoint, meth testing is done to define the extent of a building that exceeds the remediation limit. If meth activity occurs in a multifamily unit, some meth may migrate through the unit door to the exterior stairwell or interior hallway (outside the unit but inside the building), so hallway or stairwell testing is frequently performed. Sampling may be performed immediately outside the source unit’s door and can exceed a limit such as 0.5 μg/100 cm^2^. This situation triggers a need to test the hallway or stairwell again to determine how far the contamination plume migrated, and several additional tests may be needed before the plume is defined. If the plume extends to near other units, testing may be needed inside those units as well. This culminates in repeated site visits, notifications to tenants, and inevitable anxiety. In the case of condominiums, homeowner association permission is required for testing these common areas and other owners’ or tenants’ permission may be needed. When the testing is necessary, this may be justifiable, but if not, it is costly. Based on the data in 12 PAs with hallways, the average concentration was 0.6 μg/100 cm^2^, barely above the limit, and only 33% of hallways exceeded the limit. The average concentrations in the hallway were 74-fold lower than in the source unit and the contaminant plume usually did not reach other units, indicating limited migration potential. Moreover, based on the usually modest concentrations in hallways, the potential to exceed the limit in other units is also low. In summary, a remediation limit for hallways equal to the BML sometimes creates additional testing costs, inconveniences, delays, anxieties and other psychosocial costs, legal involvement, and other issues, while there is limited potential for any significant migration. The same issues may occur with exterior stairwells, which also get scarcely contaminated. These factors make it advisable to base an AML for MOAs on an appropriate increment to dose that would reduce the amount of additional testing needs.

### HVAC

It is clear that meth exposure from HVAC contamination is dissimilar to dermal and ingestion exposure in the occupied portion of HUs. The dose from surface contamination depends on dermal contact area, duration, and concentration. Both dermal contact area (cm^2^ of skin exposed) and duration of contact to the interior of HVAC is very limited and perhaps entirely insignificant. Dose calculations made under a worst-case total exposure time assumption of one hour per year indicate negligible additional dose (0.1%). This raises the question why HVAC systems require remediation to the same limits as the NOAs.

The normal process for cleaning HVAC ducts is to use a mechanical duct cleaning system (brushes, whips, etc.) connected to a HEPA-filtered vacuum system to collect the particulates re-entrained from agitation equipment. Hand cleaning with rags of the reachable sections of supply and return ducts (the terminal sections), which represent less than 5% of the total system, is then conducted. Post-remedial sampling is usually heavily or exclusively limited to terminal sections that can be reached from vents, as most state regulations do not require random sampling and because it is difficult to sample randomly selected locations in ducts (Figure 5). Thus, the majority of post-remediation sampling is not representative of contaminant concentrations throughout the ductwork, but only at the double-cleaned terminal sections. Nevertheless, it is common for the HVAC to require two or more rounds of cleaning and testing before it passes, which suggests that the inaccessible sections of ducts are likely still contaminated since they are only cleaned once and are typically not tested. Statistically representative data were not identified to compare the concentrations after remediation in random locations in duct interiors and terminal sections but in three projects the concentrations after remediation and the HU was deemed compliant were 80-fold (+/− 32) higher in the middle of ducts as compared to terminal sections. Thus, absent requirements to test random inaccessible sections of ducts, the assumption that entire duct runs are as clean as duct ends is questionable. Therefore, it is not clear if the high cost of cleaning only terminal sections is beneficial.

**Figure 5:**
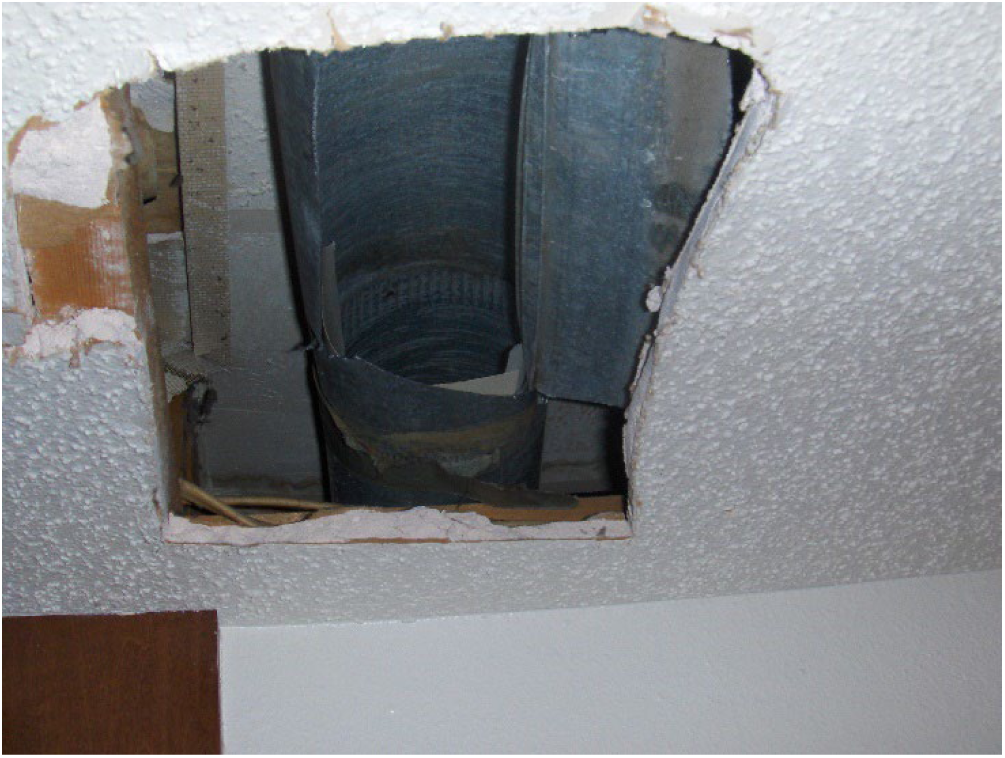
Random sampling of a supply duct in ceiling joists. Drywall had to be removed, which required asbestos testing and subsequent repairs.

The cost estimates in Figure 4 do not include some ancillary costs. HVAC systems are likely the most recalcitrant part of the house to remediate. It typically requires two additional cleanings to meet base limits (three total cleanings) and three verification samplings. Consequently, a lot of the contractor’s contingency budget is used by HVAC re-cleanings, and if the HVAC limit is increased, the contingency cost would be reduced. Also, the added cost for re-testing have not been included in the cost estimate, so an additional $500–$1,000 per project may be saved were higher limits applied and the temptation to remove and replace the system would be lower.

The actual concern with HVAC contamination is not for dermal or ingestion exposure from contaminated HVAC interior surfaces, but with potential occupant post-remediation exposure from HVAC emissions. Thus, a remediation limit for HVAC systems can be based on the concept of “potential to re-contaminate” a HU. In the 100 PAs analyzed here, for the 84 HUs with HVAC, the HVAC systems were on average 9.8-fold more contaminated than the NOAs. At some point after initial meth activity has ceased to increase HU contamination, the system would reach a quasi-equilibrium with the HU, with contaminant concentrations approximately 10 times higher than in the HU and no net efflux or influx between the system and HU. In the post-remediation datasets examined, the average meth residual in the HU was ∼0.1 μg/100 cm^2^. Based on the 10-fold higher equilibrium concentrations in HVAC, one would expect the ducts to regain a concentration of ∼1 μg/100 cm^2^ and thus, be “re-contaminated,” which could become a compliance issue upon testing at a later date. However, if a higher AML for HVAC systems were established, after remediation is finished, a new equilibrium would tend to occur with some efflux of meth into the HU. An alternative method for defining acceptable concentrations in HVAC systems is to calculate the concentration in an HVAC that would potentially add a defined amount of additional HU contamination after remediation. In a representative house, the interior surface area of the HVAC system was ∼2.5% of the surface area in the house. Given that HUs averaged 0.1 μg/100 cm^2^ in residual contamination after remediation, it may be reasonable to restrict possible recontamination from subsequent emissions for the system to a maximum of 0.2 μg/100 cm^2^. If all the meth contamination in the HVAC system was to migrate back into the house as a worst-case assumption, this criterion would not be exceeded when the HVAC system contained ≤8 μg/100 cm^2^. Thus, a remediation limit of 8 μg/100 cm^2^, 16-fold above the BML, would not lead to significant HU re-contamination. It is unlikely that all contamination would be emitted regardless; ductwork doesn’t not clean itself.

Another aspect of considering HVAC remediation limits is the choice of using composite sample results versus discrete sample results. Composite sample results are an average of the contaminant concentrations in the locations sampled. Discrete samples could be averaged to compare to remediation limits, but in Colorado and possibly some other states, the highest sample determines compliance, so the system is deemed noncompliant when any discrete sample exceeds the remediation limit even if average results do not. In 15 post-remediation sample sets, four discrete samples were collected in each HVAC system. The results ranged from 0.03–0.47 μg/100 cm^2^, with an average of 0.15 μg/100 cm^2^, compared to a BML of 0.5 μg/100 cm^2^. In effect, the remediation contractor had to re-clean the system down to a level of 0.15 μg/cm^2^ before all discrete samples were below the limit. As there is no differential exposure resulting from sections of the system with higher concentrations, and the exposure that can occur is related to the average concentration, the use of “highest discrete” result leads to over-remediation. Figure 4 provides estimates of lower failure rates and costs associated with using both the highest discrete sample versus average data.

Local exhaust systems present additional questions regarding exposure risks. The most common exhaust in HUs is the bathroom system, consisting of a louvered cover, with a fan motor behind or above it, connected to a duct that usually exits the building through a wall or roof. Bathroom exhaust fans tend to become contaminated at high levels. In the PA dataset, the average concentration in a bathroom fan system was 50 times higher than the average in the house. Similarly to HVAC systems, however, occupants rarely if ever have dermal or ingestion exposure from the system’s interior. If a composite sample is collected in a bathroom, and one aliquot is from the exhaust system, the exhaust aliquot would increase the resulting concentration by approximately 13-fold and would make the room fail even if the room itself is 13 times lower than the remediation limit. There are instances where the only area that failed in a house was the bathroom, and the bathroom only failed due to an aliquot collected from inside the fan system. Fan and duct remediation adds approximately $500–$1,000 to remediation cost, and it could be significantly more if the exhaust ducts are enclosed in drywall. A sampling professional may feel compelled to sample inside the fan so they do not “miss” a likely location that could fail or want to perform “worst-case” sampling. As the exhaust systems likely contribute very little to occupant exposure, they could simply be excluded from testing and remediation requirements. If remediation is required, it suffers the same issue as HVAC systems do, wherein they are either not sampled for compliance, or only in the terminal section that is hand cleaned. In 25 HUs, the ratio of the meth concentration in the greasy areas of the kitchen to the nearby walls was 41:1. As an organic compound with a molecular weight of 149, it is not surprising that meth would partition into organic substrates. For the same reason, other exhaust systems such as the kitchen stove range fan, attic fans, whole house fans, dryer fans, or other ventilation units could be excluded. For these reasons, testing such systems for the determination of HU compliance is misleading and may result in excess remediation costs. An alternative is to clean or remove only accessible parts of contaminated systems but excluding them from testing and from required remediation.

### Rationale for allowing incremental doses

The small incremental exposures or doses that would occur if AMLs were established for LOAs, MOAs, and HVAC systems could raise some concern as they would be “on top of” the exposure in NOAs. However, the dose occupants receive is already substantially less than the dose that corresponds to the BML. For example, in the 14 HUs in the PA dataset that were not contaminated, the average concentration in the occupied portion of the HU was 0.04 +/− 0.04 μg/100 cm^2^, less than 10% of the BML, and 99% of the occupants were exposed to concentrations of ∼0.12 μg/100 cm^2^ or less. The same is true with remediated properties. The average post-remediation NOA concentration of HUs (0.1 +/− .07 μg/100 cm^2^) indicates that over 99% of future occupants would be exposed to ∼0.3 μg/100 cm^2^ or less. This provides a margin of safety to permit appropriate increases in AMLs. All AMLs can be increased without resulting in overexposures to the majority of occupants.

A second rationale that some incremental dose is acceptable is based on the Reference Doses (RfDs) established by California and Colorado. At its remediation limit of 1.5 μg/100 cm^2^, California estimated a total dose of 0.278 μg/kg/day (at a 95% confidence interval), 8% lower than the California RfD (CalEPA 2009a, 2009b). Thus, adding AMLs that would not exceed 8% of additional dose would not exceed the RfD without even considering average real-world exposures described above. In Colorado, an even wider margin exists, as they estimated that a 0.5 μg/100 cm^2^ remediation limit would result in a dose of 0.2 μg/kg/day as compared to an RfD of 5-70 μg/kg/day. Therefore, increases in dose that fall within this range comply with the risk assessment objectives.

### Averaging of Results

In most states, any part of a property that exceeds the remediation limit is required to be mitigated. Yet, health risk is based on average exposure to occupants. An argument can be made for defining contamination under regulations based on the average concentration in the normally occupied part of HUs. In the PA dataset, 71 properties were defined as contaminated when one or more NOAs exceeded the remediation limit of 0.5 μg/100 cm^2^ (15 failed in LOAs and/or MOAs). If the average of test data were applied for the NOAs, only 54 failed, resulting in a substantial reduction in costs. However, one must consider the possibility that an occupant could spend more time in rooms that were more contaminated than the average room. This could take into account that the coefficient of variation on NOA results was ∼100%, meaning that if the average concentration in an “area” was 0.5 μg/100 cm^2^, most other areas would be all in the range of 0.5 +/− 0.5 μg/100 cm^2^.

## LIMITATIONS

This study only used contamination data from Colorado, so failure rates could vary in other states. However, use rates only vary over a four-fold range throughout the U.S. and are mostly within a factor of two-fold, and since Colorado’s use rate is close to the national average, Colorado contamination data may be a reasonable average. The cost data was obtained from contractors throughout the U.S. and cost savings would be greater or lower in other states. Some simplifications were used in determining remediation cost estimates which in general understate what the actual cost reductions would be. However, the intent was to provide a sense of costs to provide the rationale for AMLs.

## RECOMMENDATIONS

A number of recommendations ensue from this analysis. States that do not regulate meth contamination or only apply regulations to manufacturing sites may want to consider regulations on both cooking and use, considering that only a small fraction of contaminated properties are covered by use-based regulations, and there are many people occupying currently-contaminated HUs. Basing BMLs on published risk assessment values may be more appropriate than using the lowest extant limits (e.g. 0.1 and 0.05 μg/100 cm^2^), reducing costs from ∼$21K to $10K.

AMLs for LOAs should be established at a higher level such as 20-fold higher than the BML, where the incremental dose is 2.2% and the cost reduction is ∼$5K per project. AMLs for MOAs should be established at higher levels, such as two-fold. While this is a very small change in the MOA limit, it would obviate the need for a lot of additional testing, including in properties owned by others. AMLs for HVAC systems should be established at a concentration eight-fold higher than BML or more to prevent re-contamination of remediated properties. Specific numeric recommendations are outside the scope of this study, as these are risk management decisions. The purpose of the study was instead to provide a methodology for considering the issues. Local exhaust systems should be exempted from testing and remediation requirements, but remediation guidance may suggest that if they are in rooms that are contaminated and are practicable to remove or clean, optional efforts could be made in that regard. Adjustments to AMLs for HVAC, LOAs, and MOAs at once would allow remediation costs to be reduced by ∼50% or even potentially more.

## CONCLUSIONS

As meth use is increasing throughout the U.S. and more and more properties are becoming contaminated, it is appropriate to re-evaluate limits so that the remediation costs are minimized while still protecting health. The current imbalance in how remedial dollars are applied to the problem unnecessarily increases the high cost of remediation without clear benefits. Several areas where costs can be substantially reduced while still meeting the standards in published health risk assessment have been identified. Conservative limits set in some states prior to the health risk assessments publication could be raised. Minimal additional meth dose would occur if AMLs for LOAs and HVAC systems were increased several-fold, concomitantly reducing remediation costs by up to 50%. The high remediation costs have reduced the frequency of testing in many sectors, leading to more unidentified contaminated and fewer remediated properties, so increased AMLs may actually lead to improvements to environmental health.

## ACKNOWLEDGMENTS

This work was funded in part by Public Housing Authorities in Colorado. The author declares it has no competing interests but is involved in consulting activity.

## DATA AVAILABILITY STATEMENT

Supplementary data are available at 10.4121/34ce7339-6909-4e1a-9c6c-b545c4ab95de.

